# Determinants of Timely Malaria Treatment among Under-Five Children Attending Public Health Facilities in Kisumu East Sub-County, Kenya: A Health Facility-Based Cross-Sectional Study

**DOI:** 10.1101/2024.07.22.24310789

**Authors:** Geofrey Ochieng, Mutale Sampa, Patricia Maritim, Adam Silumbwe, Joseph M Zulu, Joseph Kato, Choolwe Jacobs

## Abstract

Evidence shows that most child malaria deaths occur at home shortly after symptom onset, highlighting the need for timely treatment. This study aimed to assess the determinants of timely malaria treatment among under-five children who receive care at public health facilities in Kisumu East sub-county, Kenya.

A health facility-based cross-sectional study was conducted in Kisumu East sub-county, Kenya, between 5^th^ April and 26^th^ May 2023. The study employed a two-stage stratified-cluster sampling method, first selecting hospitals and then using systematic sampling to select caregivers. Data collection was done electronically using structured questionnaires. Associations at bivariable level were assessed using either the Chi-square or Fisher’s exact test based on assumptions. Multiple logistic regression with robust standard errors was applied at a 5% significance level to establish determinants of timely malaria treatment among under-five children. STATA version 16 (College Station, Texas 77845 USA) was used for all analyses.

The sample included a total of 434 caregivers of under-five children. The study revealed that caregivers’ ability to recognize malaria symptoms was associated with higher odds of seeking timely malaria treatment for their children (AOR=2.92; 95% CI=1.36-6.25; p=0.006). Additionally, having health insurance cover was associated with higher odds of timely treatment (AOR=2.12; 95% CI=1.25-3.59; p=0.005). Those who visited herbalists before seeking care were less likely to seek timely treatment (AOR=0.13; 95% CI=0.05-0.34; p<0.0001). Fear of drugs’ side effects reduced the odds of timely treatment (AOR=0.50; 95% CI=0.29-0.87; p=0.013).

Timely malaria treatment was linked to the ability to tell symptoms and having health insurance, while delayed malaria treatment was related to herbalist visits and fear of malaria, among others. There is need for relevant stakeholders to implement strategies that address misconceptions about drug side effects, offer affordable insurance, integrate the informal health sector, and educate caregivers about under-five malaria symptoms in order to promote timely malaria treatment.

## Background

Malaria is one of the world’s major public health problems and is a common leading cause of morbidity and mortality in developing countries, despite being preventable and treatable (1). In 2021, about 4 billion people were at risk of malaria infection worldwide (1), and 247 million new cases and 619,000 deaths were recorded in 2021(2). About 95% of the global deaths were from the African continent with 78% of these deaths occurring among children under the age of five (3). Malaria continues to be a major health concern in Kenya, representing 13% to 15% of outpatient consultations (4). Annually, the country experiences around 3.5 million new cases and 10,700 deaths, with the western region, including Kisumu sub-county, being a high-risk area (5).

Timely malaria treatment is defined as early diagnosis and prompt treatment using appropriate antimalarial within 24 hours of onset of symptoms (1). Kenya embraced timely malaria treatment as an intervention in 2000 as part of the adoption of the Abuja declaration (6,7). The initiative received further support in 2007 with the implementation of the President’s Malaria Initiative, which focused on enhancing malaria diagnostics, effective treatment, and strengthening supply chain management (6,7). The Kenyan government has undertaken various strategies to promote timely treatment such as training private medicine retailers for effective case management and engaging community health volunteers (CHVs) in the distribution of anti-malarial drugs (8,9). In further efforts to promote timely malaria treatment, Kenya eliminated user fees for children under five with malaria in 2004, introducing a minimal flat registration fee of Kenya shillings10 to Kenya shillings 20 (USD 0.08 to USD 0.15) at public dispensaries and hospitals (10) and further set a goal to treat 90% of childhood fevers with effective anti-malarial medicines within 24 hours of symptom onset by 2023 (8).

Evidence shows that most of the malaria deaths occur at home without receiving proper medical care, and in case care is sought, it is often delayed (11,12). Several studies have investigated the determinants influencing the timely treatment of malaria among children under five children and have unveiled a spectrum of predictors of timely malaria treatment spanning socio-cultural, physical, and economic dimensions (13–27). Despite all these efforts, the proportion of under-five children in Kisumu East Sub-county receiving timely malaria treatment remains low as evidenced in Kenya Malaria Indicator Survey (KMIS) of 2020 which reported that only 36% of children with recent fever received timely treatment on the same or next day following fever onset (4). Additionally, while studies have investigated the determinants of timely malaria treatment, there are limited studies concerning this health issue in Kisumu East sub-county. Moreover, the existing studies have produced conflicting findings (13–27), adding a layer of complexity to the understanding of the factors influencing timely malaria treatment. For example, studies done in Northwest Ethiopia showed that malaria knowledge was a significant determinant of malaria treatment (20,23) while another study done in southwest Ethiopia showed that malaria knowledge was not a significant determinant of timely malaria treatment among under-five. These discrepancies are attributed to varying socio-cultural and socio-economic factors that are context-specific (25,28).

Therefore, for the Kenya to meet its set target, there is need to understand and explore the determinants of timely malaria treatment among under-five in this context. The results from this study could help inform/strengthen the implementation future interventions, increasing timely malaria treatment in under-fives and aligning with SDG 3.2 for preventing child malaria deaths. This study therefore sought to identify determinants of timely treatment among under-five children suffering from malaria in Kisumu East Sub-County, Kenya.

## Methods

### Conceptual framework

This study employed Levesque et al Conceptual Framework for Patient-centred access to health to define access to timely malaria treatment. This framework defines access comprehensively as the opportunity to identify healthcare needs, to seek healthcare services, to reach, to obtain or use health care services and to actually have the need for services fulfilled (29). This conceptual framework bridges supply-side aspects of healthcare systems and organizations with demand-side characteristics of populations, while also considering the processes involved in realizing access (29). The five supply-side components of access to healthcare according to this framework include approachability, acceptability, availability and accommodation, affordability, and appropriateness while demand-side components of access to healthcare include the ability to perceive, the ability to seek, the ability to reach, the ability to pay, and the ability to engage (29). The study primarily focused on demand-side determinants influencing timely malaria treatment among under-five children.

### Study design

A health facility-based cross-sectional study design was conducted to assess the determinants of timely treatment among under-five children seeking care for malaria treatment in from public health facilities in Kisumu East sub-county, Kenya. This design was chosen for its relevance to this study’s objectives. Additionally, hospitals serve as key access points for healthcare services, especially for under-five children seeking malaria. The design is also resource-efficient and can be conducted within a reasonable timeframe, which made it well-suited for addressing the research question in this study within the constraints of time and resources that were available.

### Study setting

This study was conducted in Kisumu East sub-county in Kisumu County in Western Kenya between April 2023 and May 2023. Kisumu East sub-county is one of the six sub-counties in Kisumu County in Kenya. Based on data from the 2019 census by the Kenya Bureau of Statistics (KNBS), it has a population of 220,997, with 51% of the population being female (30). The sub-county has 5 wards namely Kajulu, Kolwa East, Manyatta B, Nyalenda A, and Kolwa central and covers 142 square kilometres. Given it boarders the lake, all the wards in the sub-county are malaria endemic. Kisumu East sub-county was selected due to its location in the western region, where the percentage of under-five children receiving care or advice from healthcare providers for fever dropped to 57% in 2020 from 66% in 2015 (4).

### Study population

The study population consisted of caregivers/parents of under-five children suffering from malaria who sought malaria treatment in the public health facilities between 5^th^ April and 26^th^ May 2023. It included caregivers/parents of under-five with malaria who presented to the 7 seven public health facilities of study to seek malaria treatment and confirmed by microscopy or RDT for *Plasmodium* species. Caregivers of children who had sought treatment elsewhere before coming to the designated health facilities were excluded, as were caregivers under 18 and were not the sole caregivers in child-headed households.

### Sample size determination

A sample of 446 caregivers was determined using a cross-sectional sample size formula in Epi Info version 7.2, with parameters set at a 95% confidence level, 80% power, and a 5% margin of error. The proportion of under-five children receiving timely treatment, as reported by the Kenya Malaria Indicator Survey (KMIS) 2021 (36%), informed the calculation (KMIS, 2021). To account for non-response and clustering, a 5% non-response rate and a design effect of 1.2 were incorporated, based on statistics from the Kenya Malaria Indicator Survey of 2020 (4).

### Sampling technique and procedure

A two-stage stratified cluster sampling method was used to select participants. Kisumu East sub-county was divided into rural and urban strata, with all 14 public health facilities serving as clusters. In the first stage, 7 health facilities were chosen using a probability proportional to size (PPS) based on the number of confirmed malaria cases in under-five children from sub-county data. In the second stage, systematic sampling with a systematic sampling interval (k) of 1 was employed, resulting in 64 caregivers selected at each public health facilities selected in the first stage.

### Study variables

The outcome variable was timely malaria treatment and was defined as early diagnosis and prompt treatment using appropriate antimalarial within 24 hours of onset of symptoms (1). Timely malaria treatment referred to caregivers of under-five with malaria who presented to a public health facility to seek malaria treatment within 24 hours of the onset of the symptoms and confirmed by microscopy or RDT for *Plasmodium* species Delayed treatment indicated caregivers/parents seeking treatment after 24 hours of symptom onset, similarly, confirmed by microscopy or RDT for *Plasmodium* species. The study considered various explanatory variables, including demographic factors such as the age of the child and caregiver, residence, education level, and the child’s sex. Socio-cultural factors encompassed malaria knowledge, the ability to recognize symptoms, belief in using approved malaria drugs, trust in healthcare workers, herbalist visits, over-the-counter medication use, gender of the household head, and decision-making authority for seeking care. Physical factors involved the time taken to reach a health facility, transportation accessibility, and the mode of transportation. Additionally, socio-economic and engagement factors comprised household income, cost of care, health insurance, social support, history of timely care, history of child’s death, and caregiver support to other caregivers. Malaria knowledge was assessed through six knowledge questions, with each correct response earning a participant a score of 1. The composite score was calculated and converted into a percentage for comparison with Bloom’s cut-off points. Participants scoring 80.0–100.0% demonstrated good knowledge, while those with scores of 60.0–79.0% had satisfactory knowledge. A score below 60.0% indicated poor knowledge.

### Data collection tool and procedure

A structured questionnaire, adapted from prior studies on a similar topic (13,14,20–22,27,31,32), was administered to participants by trained community health volunteers (CHVs). The questionnaire was initially prepared in English, translated into the local language, Luo, and then back translated to English for consistency. Caregivers from different health facilities were interviewed, with informed consent obtained before the interviews. After under-five children had been diagnosed by a clinician/doctor at the health facility, caregivers were interviewed in face-to-face exit interviews by CHVs using Computer Assisted Personal Interviews (CAPI), specifically Kobo Collect.

### Data quality control

Before data collection, a pre-test of the questionnaire was carried out to identify any necessary adjustments. Fourteen participants from two health facilities that were not part of the study were involved in the pre-test. Comprehensive one-day training on questionnaire completion was provided to the CHVs, and during data collection, at the end of each day, the questionnaires were reviewed for completeness before submission to the server.

### Data management and analysis

Data was directly exported to STATA version 16 (College Station, Texas 77845 USA) from the server for analysis. Recoding of variables was done, and composite variables were generated. Range and consistency checks were performed, ensuring dataset completeness. Descriptive statistics involved frequency distributions for categorical variables and summary statistics for continuous variables. Continuous variables, namely the age of caregivers and under-five children, underwent normality testing using the Shapiro-Wilk test and were confirmed through probability plots. It was determined that both variables exhibited skewness, leading to the reporting of their median values and interquartile ranges.

Bivariable analysis aimed to identify potential variables associated with timely malaria treatment for inclusion in the multivariable analysis. Dependent on the assumptions, the chi-square test or Fisher’s chi-square test was utilized at a 20% level of significance to assess the association between timely malaria treatment and each categorical explanatory variable. The choice between the chi-square test and Fisher’s exact test depended on whether the chi-square assumptions were met. Simple logistic regression was employed to identify potential continuous explanatory variables at a 20% significance level. Higher level of significance was chosen to allow more explanatory variables to be included at multivariable analysis stage.

In the multivariable analysis, a multiple logistic regression model with robust standard errors was employed to identify the determinants of timely malaria treatment at a 95% confidence interval. The standard multiple logistic regression model was considered inappropriate due to potential bias in standard errors resulting from the clustering effect within the sampling design (33). Hence, a multiple logistic regression model with robust standard errors was used to account for this. All significant variables identified at the bivariable level were included in the model, and an investigator-led stepwise regression approach was applied. To determine the best-fit model, the Akaike Information Criteria (AIC) and the Bayesian Information Criteria (BIC) were used. Multicollinearity was tested using variance inflation factor (VIF) and a value less than 10 was included in the model. Performance of the model was checked using sensitivity and specificity, percent of correct specification and area under Receiver Operating Characteristic Curve (ROC).

### Ethical considerations

Ethical approval for this study was obtained from the University of Zambia Biomedical Research Ethics Committee (reference number 3446-2022). Additionally, another ethical clearance was obtained from the Maseno University Ethics Review Committee (reference number MSU/DRPI/MUSERC/01189/23). A research permit was also secured from the National Commission for Science, Technology, and Innovation with license number NACOSTI/P/23/24218. Caregivers interviewed in the study provided written consent for their participation. Furthermore, as our study included minors, we made sure to we obtained verbal consent from the caregivers of under-five children suffering from malaria. The consent process was documented and witnessed by the health facility in-charges at each of the different hospitals.

## Results

### Study Population Characteristics

In this study, a total of 434 caregivers were included, with each of the seven health facilities contributing 62 caregivers. The age of caregivers included in this study was not normally distributed and had a median of 27 (Interquartile range (IQR): 24 32) years. Similarly, age of the under-five children was skewed with a median of 16 (IQR: 8 30) months and most of the under-fives were males accounting to 52.53% (228/434). Furthermore, 43.03% (187/434) of the caregivers attained a secondary education, 41.01% (178/434) had a primary school education, 13.82% (60/434) had at least a college education while only 2.08% (9/434) had no education. A substantial majority of caregivers 74.88% (325/434) had a household income of less than ksh 10,000. A minority of the caregivers, 31.80% (138/434) had insurance cover. Regarding caregiver support, majority of caregivers representing 97.70% (424/434), reported having support from the healthcare workers. Table 1 summarizes the Study Population Characteristics.

**Table 1:**
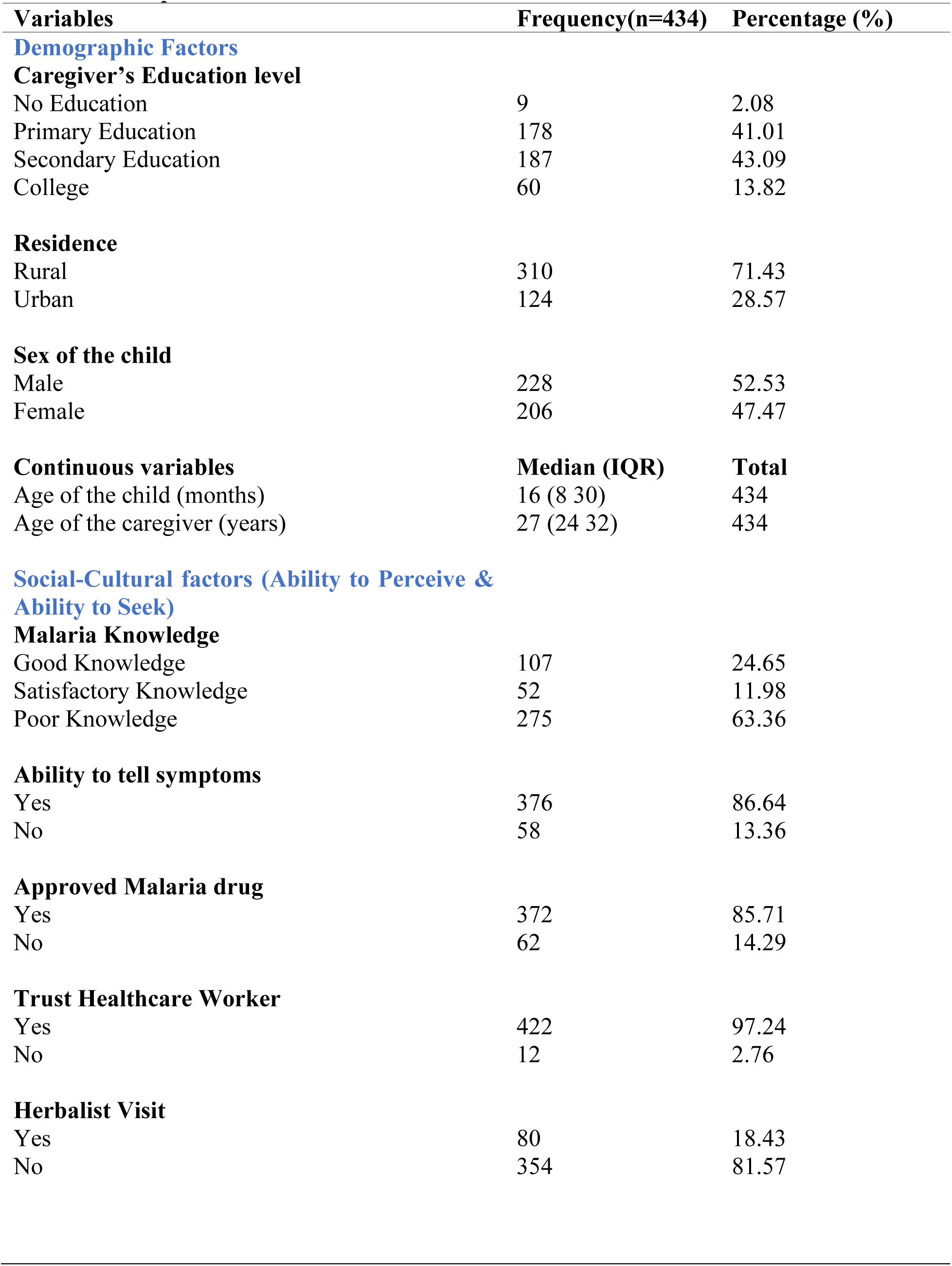
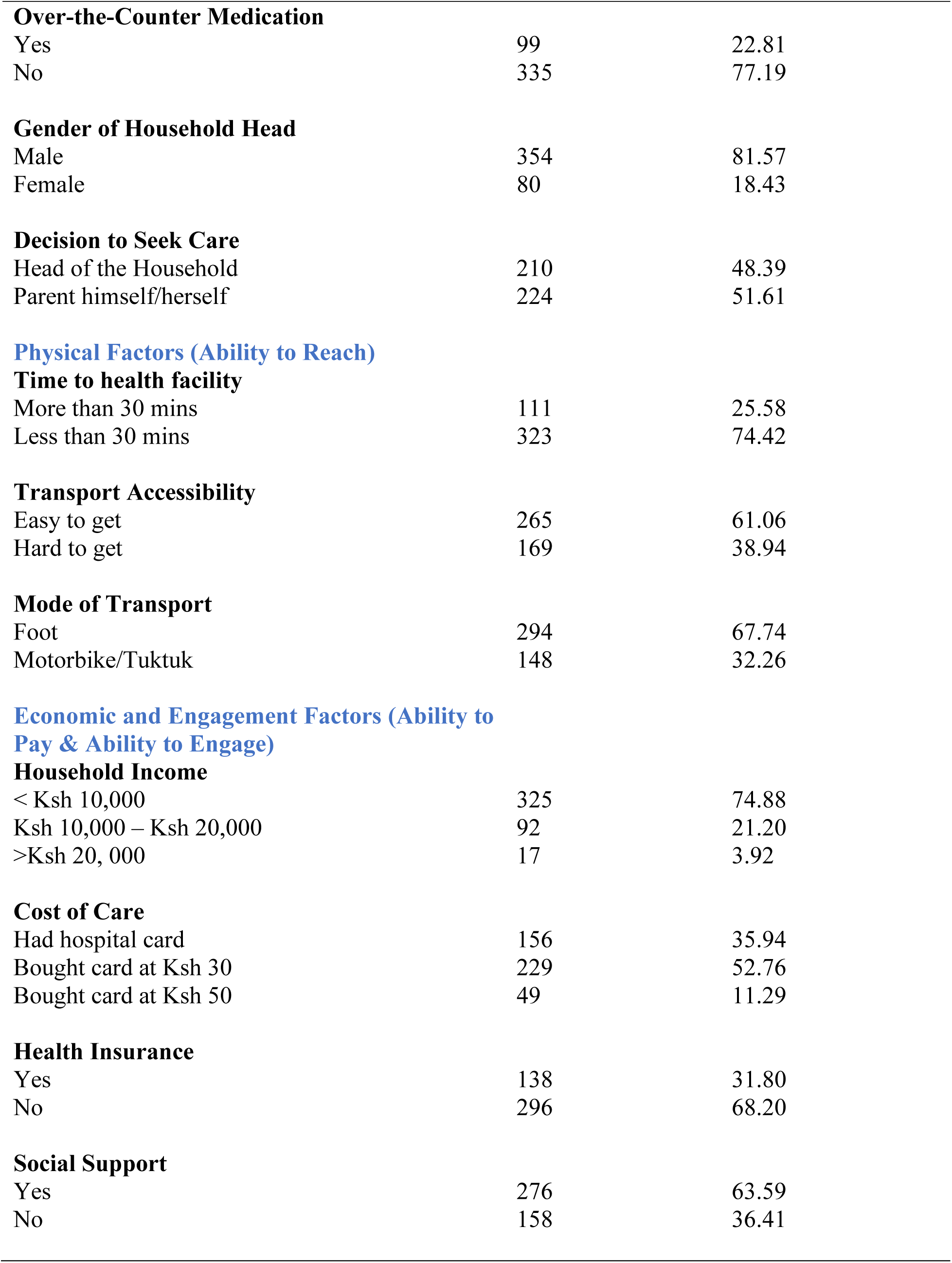
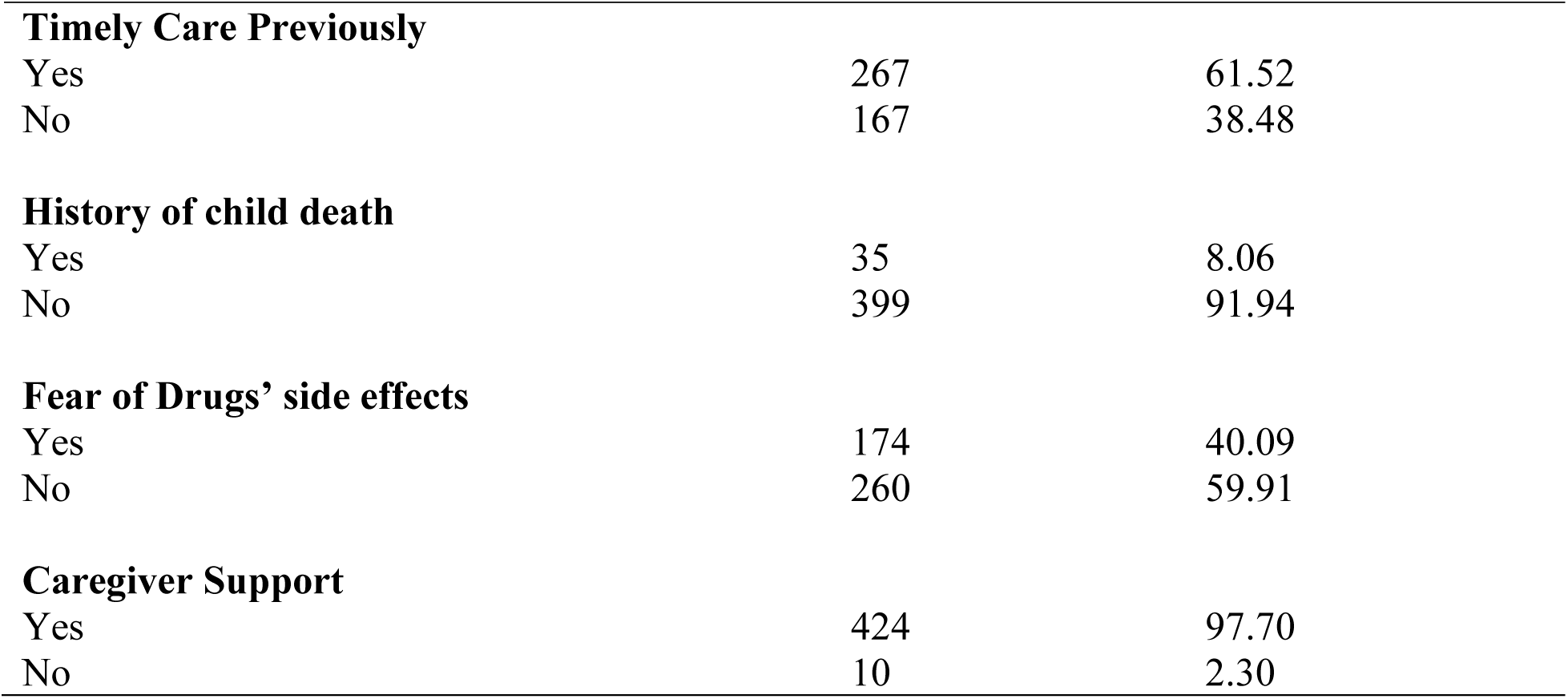
Descriptive Statistics.

### Knowledge and health seeking practices

Most of the caregivers 63.36% (275/434) poor knowledge about malaria while only 24.65% (107/434) had good knowledge with 11.98% (52/434) having satisfactory knowledge. Most caregivers, 86.64% (376/434) were able to identify malaria symptoms in their under-five. Additionally, most caregivers’ households were male headed accounting 81.57% (354/434). A larger proportion of caregivers, 86.64% (376/434) reported being able to tell when their children started showing symptoms. Furthermore, only 18.43% (80/434) sought treatment from herbalist or traditional healer before presenting to health facility. Regarding physical factors, caregivers primarily utilized two modes of transport for seeking medical care for their children which were foot and motorbike/tuktuk. Most of the caregivers came to the health facility by foot accounting to 67.74% (294/434) while 32.26% (148/434) used motorbike/tuktuk as their mode of transportation to the health facility. Majority of caregivers, 74.42% (323 /434), took less than 30 minutes to reach the health facility.

A significant proportion of caregivers 60.14% (261/434) sought malaria treatment timely (within 24 hours) for their sick under-five children. Delayed care seeking (after 24 hours) for malaria was observed among 39.86% (173/434) of participants. Figure 1 represents the distribution of how caregivers sought treatment.

**Figure 1:**
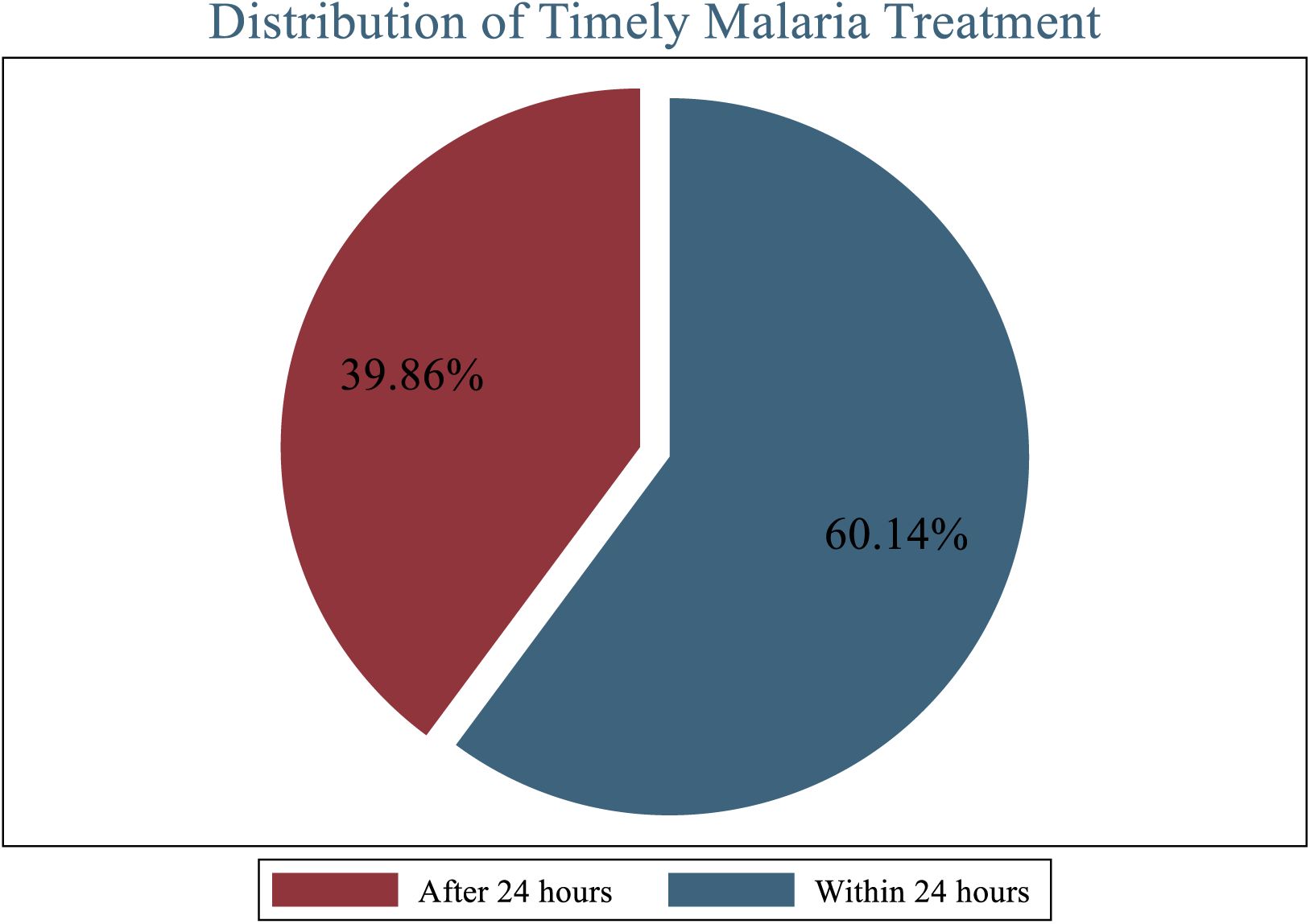
Distribution of Malaria Treatment Seeking by Caregivers

### Determinants of timely malaria treatment

According to bivariable analysis, the results from of chi-square test indicated various factors significantly associated with timely malaria treatment at a significance level of 20%. These variables included the ability of caregivers to tell symptoms (p-value<0.0001), malaria knowledge (p-value =0.022), use of approved malaria drugs (p-value<0.0001), herbalist visits (p-value<0.0001), use of over-the-counter medication (p-value=0.001), gender of the household head (p-value =0.040), decision-maker for seeking care (p-value=0.001), cost of care (p-value<0.0001), health insurance (p-value<0.0001), social support (p-value=0.001), previous timely care (p-value<0.0001), fear of drugs’ side effects (p-value<0.0001), and residence (p-value=0.038) (Table 2). Conversely, from the Fisher’s exact test, the educational level of caregivers was significantly associated with timely malaria treatment (p-value=0.195) (Table 2). For simple logistic regression, age of the caregivers was significant (p-value= 0.008) (Table 2). **[Table 2]**

**Table 2.**
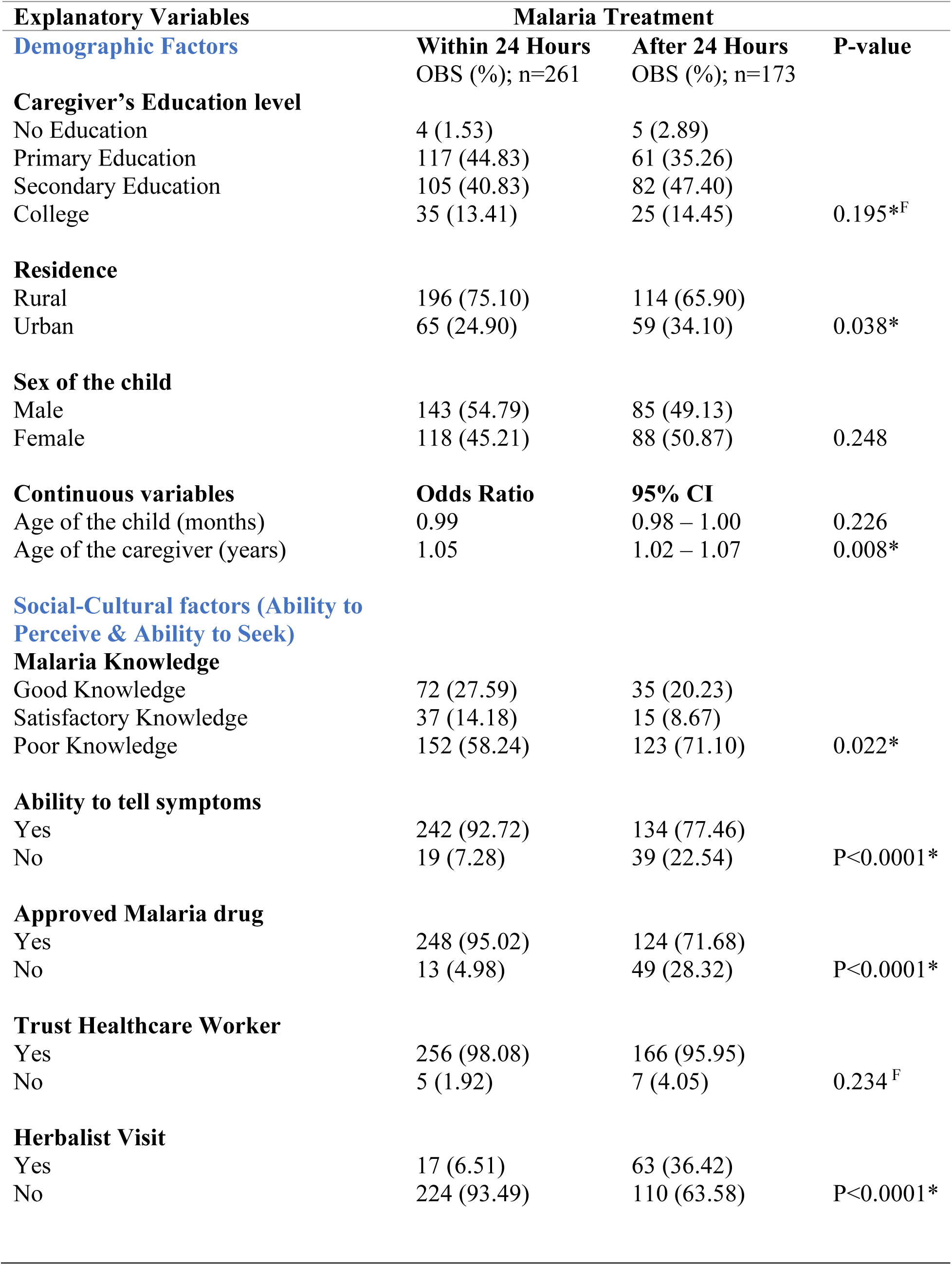
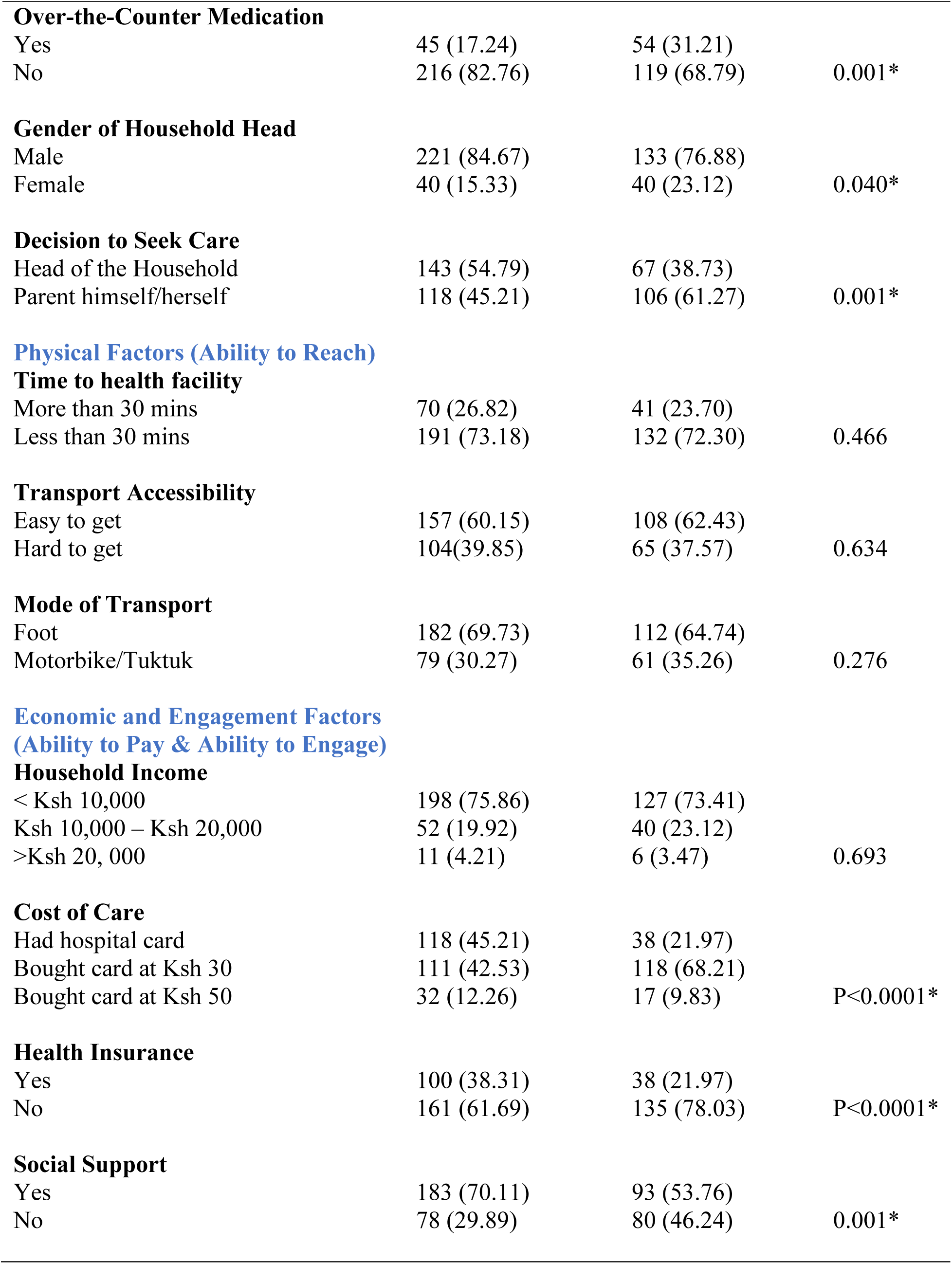
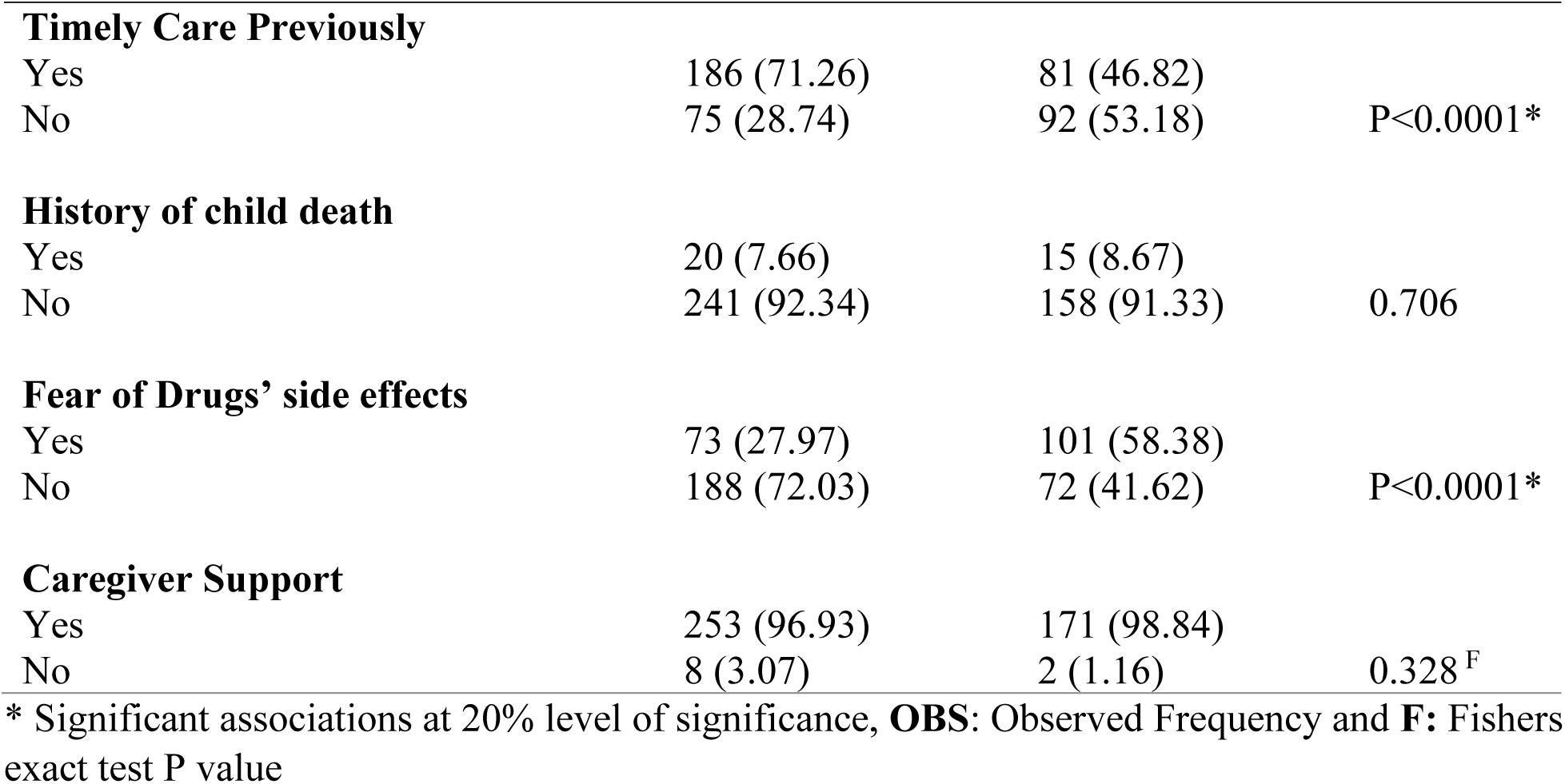
Factors associated with timely malaria treatment (Bivariable analysis results)

| **Explanatory Variables** | **Malaria Treatment** |  |  |
| --- | --- | --- | --- |
| **Demographic Factors** | **Within 24 Hours** | **After 24 Hours** | **P-value** |
|  | OBS (%); n=261 | OBS (%); n=173 |  |
| **Caregiver's Education level** |  |  |  |
| No Education | 4 (1.53) | 5 (2.89) |  |
| Primary Education | 117 (44.83) | 61 (35.26) |  |
| Secondary Education | 105 (40.83) | 82 (47.40) |  |
| College | 35 (13.41) | 25 (14.45) | 0.195*<sup>F</sup> |
| **Residence** |  |  |  |
| Rural | 196 (75.10) | 114 (65.90) |  |
| Urban | 65 (24.90) | 59 (34.10) | 0.038* |
| **Sex of the child** |  |  |  |
| Male | 143 (54.79) | 85 (49.13) |  |
| Female | 118 (45.21) | 88 (50.87) | 0.248 |
| **Continuous variables** | **Odds Ratio** | **95% CI** |  |
| Age of the child (months) | 0.99 | 0.98 – 1.00 | 0.226 |
| Age of the caregiver (years) | 1.05 | 1.02 – 1.07 | 0.008* |
| **Social-Cultural factors (Ability to Perceive & Ability to Seek)** |  |  |  |
| **Malaria Knowledge** |  |  |  |
| Good Knowledge | 72 (27.59) | 35 (20.23) |  |
| Satisfactory Knowledge | 37 (14.18) | 15 (8.67) |  |
| Poor Knowledge | 152 (58.24) | 123 (71.10) | 0.022* |
| **Ability to tell symptoms** |  |  |  |
| Yes | 242 (92.72) | 134 (77.46) |  |
| No | 19 (7.28) | 39 (22.54) | P<0.0001* |
| **Approved Malaria drug** |  |  |  |
| Yes | 248 (95.02) | 124 (71.68) |  |
| No | 13 (4.98) | 49 (28.32) | P<0.0001* |
| **Trust Healthcare Worker** |  |  |  |
| Yes | 256 (98.08) | 166 (95.95) |  |
| No | 5 (1.92) | 7 (4.05) | 0.234<sup>F</sup> |
| **Herbalist Visit** |  |  |  |
| Yes | 17 (6.51) | 63 (36.42) |  |
| No | 224 (93.49) | 110 (63.58) | P<0.0001* |
| **Over-the-Counter Medication** |  |  |  |
| Yes | 45 (17.24) | 54 (31.21) | 0.001* |
| No | 216 (82.76) | 119 (68.79) |  |
| **Gender of Household Head** |  |  |  |
| Male | 221 (84.67) | 133 (76.88) | 0.040* |
| Female | 40 (15.33) | 40 (23.12) |  |
| **Decision to Seek Care** |  |  |  |
| Head of the Household | 143 (54.79) | 67 (38.73) | 0.001* |
| Parent himself/herself | 118 (45.21) | 106 (61.27) |  |
| **Physical Factors (Ability to Reach)****Time to health facility** |  |  |  |
| More than 30 mins | 70 (26.82) | 41 (23.70) | 0.466 |
| Less than 30 mins | 191 (73.18) | 132 (72.30) |  |
| **Transport Accessibility** |  |  |  |
| Easy to get | 157 (60.15) | 108 (62.43) | 0.634 |
| Hard to get | 104(39.85) | 65 (37.57) |  |
| **Mode of Transport** |  |  |  |
| Foot | 182 (69.73) | 112 (64.74) | 0.276 |
| Motorbike/Tuktuk | 79 (30.27) | 61 (35.26) |  |
| < Ksh 10,000 | 198 (75.86) | 127 (73.41) | 0.693 |
| Ksh 10,000 – Ksh 20,000 | 52 (19.92) | 40 (23.12) |  |
| >Ksh 20, 000 | 11 (4.21) | 6 (3.47) |  |
| **Cost of Care** |  |  |  |
| Had hospital card | 118 (45.21) | 38 (21.97) | P<0.0001* |
| Bought card at Ksh 30 | 111 (42.53) | 118 (68.21) |  |
| Bought card at Ksh 50 | 32 (12.26) | 17 (9.83) |  |
| **Health Insurance** |  |  |  |
| Yes | 100 (38.31) | 38 (21.97) | P<0.0001* |
| No | 161 (61.69) | 135 (78.03) |  |
| **Social Support** |  |  |  |
| Yes | 183 (70.11) | 93 (53.76) | 0.001* |
| No | 78 (29.89) | 80 (46.24) |  |

| *Table 2 Continued* |  |  |  |
| --- | --- | --- | --- |
| **Timely Care Previously** |  |  |  |
| Yes | 186 (71.26) | 81 (46.82) | P<0.0001* |
| No | 75 (28.74) | 92 (53.18) |  |
| **History of child death** |  |  |  |
| Yes | 20 (7.66) | 15 (8.67) | 0.706 |
| No | 241 (92.34) | 158 (91.33) |  |
| **Fear of Drugs' side effects** |  |  |  |
| Yes | 73 (27.97) | 101 (58.38) | P<0.0001* |
| No | 188 (72.03) | 72 (41.62) |  |
| **Caregiver Support** |  |  |  |
| Yes | 253 (96.93) | 171 (98.84) | 0.328<sup>F</sup> |
| No | 8 (3.07) | 2 (1.16) |  |

\* Significant associations at 20% level of significance, **OBS**: Observed Frequency and **F**: Fishers exact test P value

Caregivers who could accurately identify when their children began showing symptoms of malaria were 2.92 times more likely to seek treatment within 24 hours for their sick under-five children (AOR=2.92, 95% CI=1.36–6.25, p-value=0.006) compared to those who were those that were unable able to tell when their children started showing symptom(s) (Table 3). Caregivers who visited herbalist or traditional healer before seeking care were 0.13 times less likely to seek timely malaria treatment compared to those caregivers who did not visit herbalist or traditional healer before seeking care (AOR = 0.13; 95% CI = 0.05 – 0.34; p-value < 0.0001). Caregivers who administered over-the-counter medication at home before consulting a healthcare provider were 6.51 times more likely to seek timely malaria treatment for their children under five years old, compared to caregivers who did not utilize over-the-counter medication before visiting a health facility (AOR=6.51; 95% CI= 2.83 – 14.97; p-value<0.0001) (Table 3). Households where decision to seek care was made by caregivers themselves, the under-five children were 0.44 times less likely to receive malaria treatment within 24 hours as compared to households where decision to seek care was made by the household heads (AOR=0.44; 95% CI= 0.23 – 0.76; p-value=0.001) (Table 3).

**Table 3:**
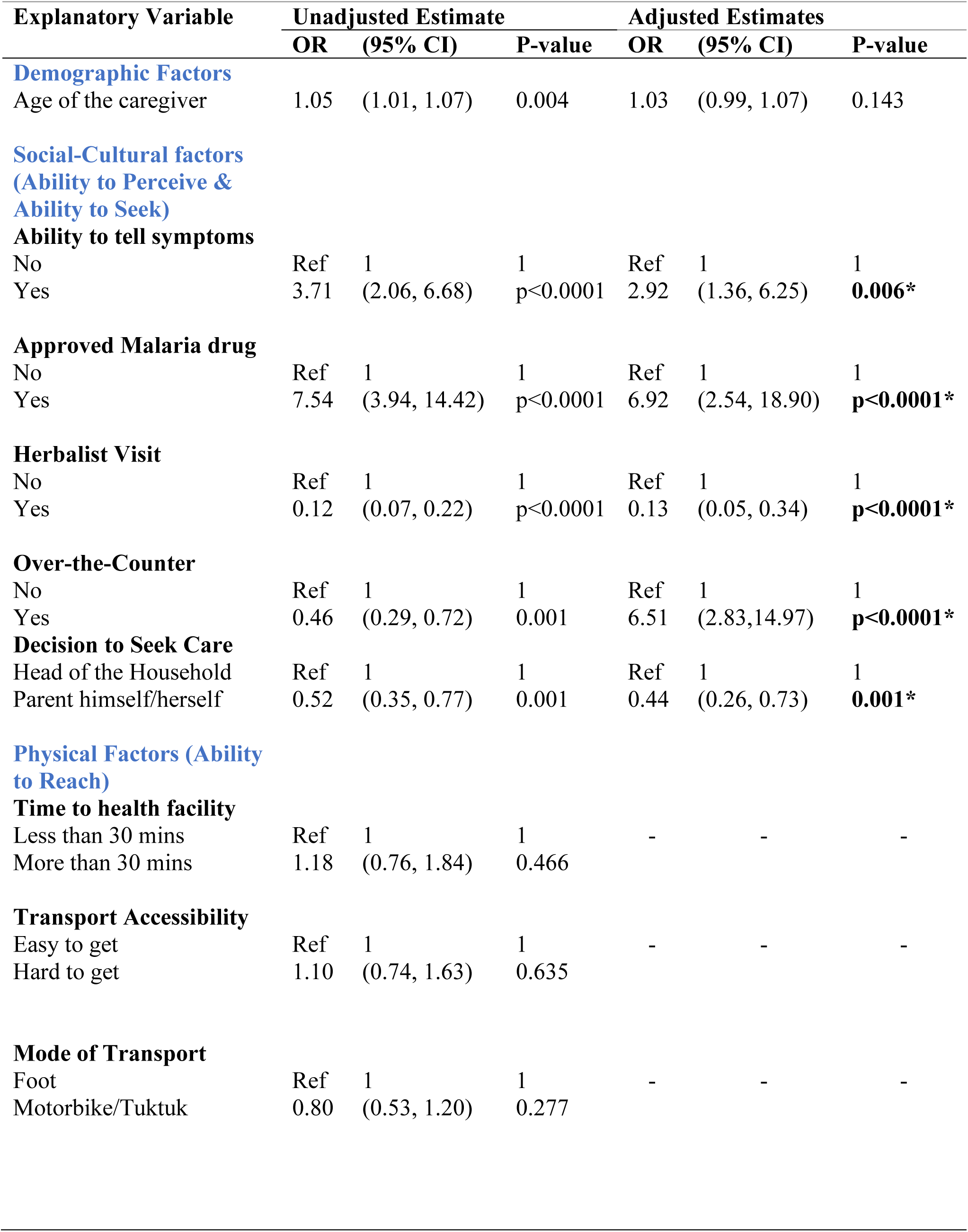
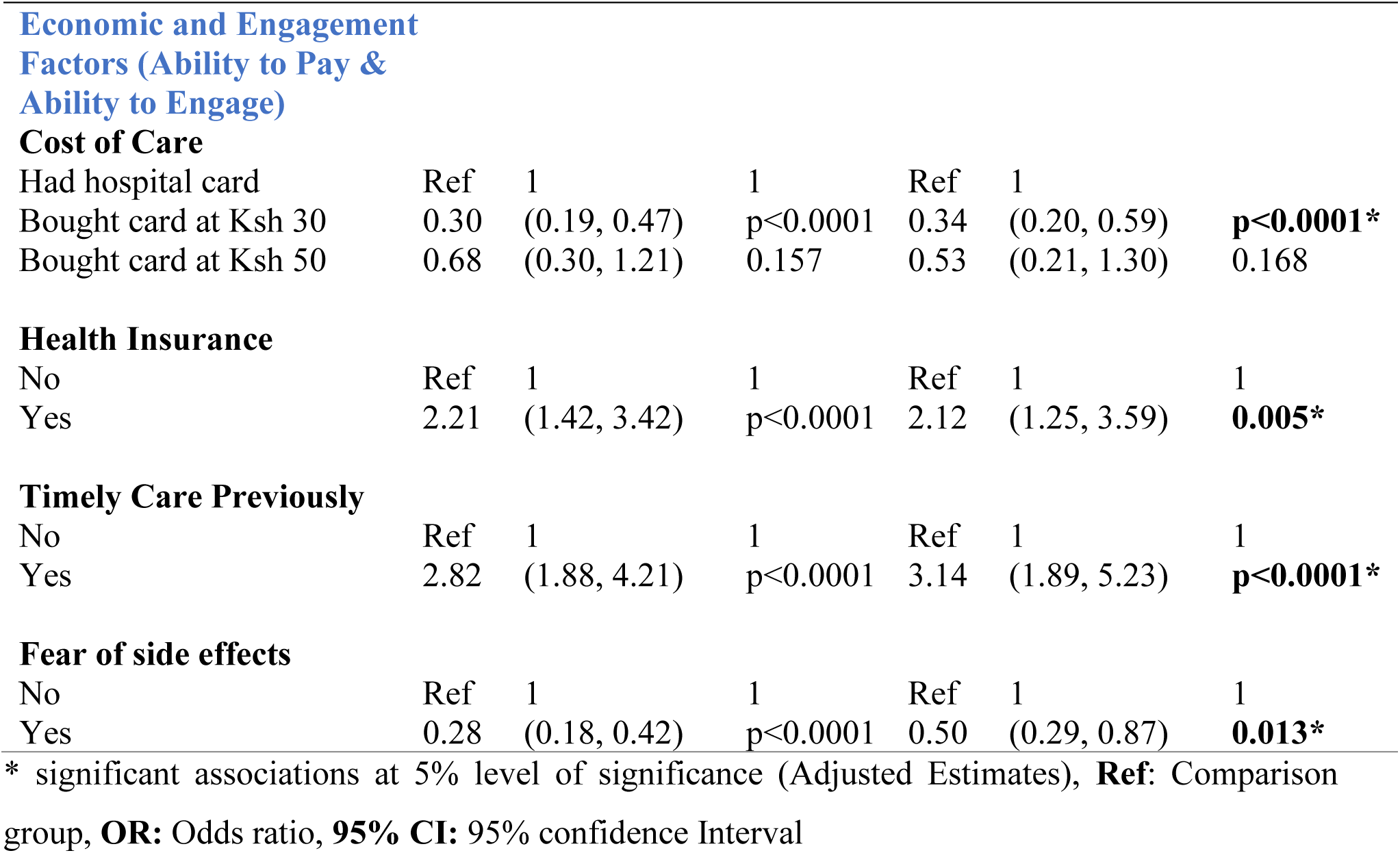
Factors associated with timely malaria treatment among under-fives.

| Explanatory Variable | Unadjusted Estimate |  |  | Adjusted Estimates |  |  |
| --- | --- | --- | --- | --- | --- | --- |
|  | OR | (95% CI) | P-value | OR | (95% CI) | P-value |
| **Demographic Factors** |  |  |  |  |  |  |
| Age of the caregiver | 1.05 | (1.01, 1.07) | 0.004 | 1.03 | (0.99, 1.07) | 0.143 |
| **Social-Cultural factors**<br>**(Ability to Perceive &**<br>**Ability to Seek)** |  |  |  |  |  |  |
| **Ability to tell symptoms** |  |  |  |  |  |  |
| No | Ref | 1 | 1 | Ref | 1 | 1 |
| Yes | 3.71 | (2.06, 6.68) | p<0.0001 | 2.92 | (1.36, 6.25) | **0.006\*** |
| **Approved Malaria drug** |  |  |  |  |  |  |
| No | Ref | 1 | 1 | Ref | 1 | 1 |
| Yes | 7.54 | (3.94, 14.42) | p<0.0001 | 6.92 | (2.54, 18.90) | **p<0.0001\*** |
| **Herbalist Visit** |  |  |  |  |  |  |
| No | Ref | 1 | 1 | Ref | 1 | 1 |
| Yes | 0.12 | (0.07, 0.22) | p<0.0001 | 0.13 | (0.05, 0.34) | **p<0.0001\*** |
| **Over-the-Counter** |  |  |  |  |  |  |
| No | Ref | 1 | 1 | Ref | 1 | 1 |
| Yes | 0.46 | (0.29, 0.72) | 0.001 | 6.51 | (2.83,14.97) | **p<0.0001\*** |
| **Decision to Seek Care** |  |  |  |  |  |  |
| Head of the Household | Ref | 1 | 1 | Ref | 1 | 1 |
| Parent himself/herself | 0.52 | (0.35, 0.77) | 0.001 | 0.44 | (0.26, 0.73) | **0.001\*** |
| **Physical Factors (Ability**<br>**to Reach)** |  |  |  |  |  |  |
| **Time to health facility** |  |  |  |  |  |  |
| Less than 30 mins | Ref | 1 | 1 | - | - | - |
| More than 30 mins | 1.18 | (0.76, 1.84) | 0.466 |  |  |  |
| **Transport Accessibility** |  |  |  |  |  |  |
| Easy to get | Ref | 1 | 1 | - | - | - |
| Hard to get | 1.10 | (0.74, 1.63) | 0.635 |  |  |  |
| **Mode of Transport** |  |  |  |  |  |  |
| Foot | Ref | 1 | 1 | - | - | - |
| Motorbike/Tuktuk | 0.80 | (0.53, 1.20) | 0.277 |  |  |  |
| Had hospital card | Ref | 1 | 1 | Ref | 1 |  |
| Bought card at Ksh 30 | 0.30 | (0.19, 0.47) | p<0.0001 | 0.34 | (0.20, 0.59) | **p<0.0001\*** |
| Bought card at Ksh 50 | 0.68 | (0.30, 1.21) | 0.157 | 0.53 | (0.21, 1.30) | 0.168 |
| **Health Insurance** |  |  |  |  |  |  |
| No | Ref | 1 | 1 | Ref | 1 | 1 |
| Yes | 2.21 | (1.42, 3.42) | p<0.0001 | 2.12 | (1.25, 3.59) | **0.005\*** |
| **Timely Care Previously** |  |  |  |  |  |  |
| No | Ref | 1 | 1 | Ref | 1 | 1 |
| Yes | 2.82 | (1.88, 4.21) | p<0.0001 | 3.14 | (1.89, 5.23) | **p<0.0001\*** |
| **Fear of side effects** |  |  |  |  |  |  |
| No | Ref | 1 | 1 | Ref | 1 | 1 |
| Yes | 0.28 | (0.18, 0.42) | p<0.0001 | 0.50 | (0.29, 0.87) | **0.013\*** |

\* significant associations at 5% level of significance (Adjusted Estimates), **Ref:** Comparison group, **OR:** Odds ratio, **95% CI:** 95% confidence Interval

The study also revealed that caregivers who had health insurance cover were 2.12 times more likely to seek timely malaria treatment compared to those without health insurance cover (AOR= 2.12; 95% CI = 1.25 – 3.59; p-value = 0.005) (Table 3). Caregivers who had previously sought timely treatment for their under-five children were 3.14 times more likely to seek timely malaria treatment again compared to caregivers who had never sought timely treatment before (AOR=3.14; 95% CI=1.89 – 5.23; p-value<0.0001). Lastly, caregivers who expressed fear of side effects in their under-five children were 0.50 times less likely to seek timely malaria treatment compared to those without such fears (AOR= 0.50, 95% CI = 0.29 – 0.87, p-value = 0.013). The study however, did not reveal any statistically significant associations between physical factors and timely malaria treatment among under-five children.

## Discussion

This study aimed at assessing determinants of timely malaria treatment among under-five children attending public health facilities in Kisumu East sub-county in Kenya. Utilizing the Levesque’s conceptual Framework for Patient-Centered Access to Health, the study identified that four of the five patient abilities (perceive, seek, pay, and engage) significantly influenced timely malaria treatment in this population. Regarding the ability to perceive, caregivers who demonstrated the capability to recognize malaria symptoms in children under five were more likely to seek timely malaria treatment for their under-five. This finding was consistent with a similar study conducted in Mali, Nigeria, and Madagascar (18). Regarding ability to pay, having health insurance was linked to a higher likelihood of timely care for under-five children with malaria, which was consistent with the findings of a similar study in Guinea (34). In terms of ability to seek, it was observed that seeking treatment from traditional healers prior to presenting to formal health facilities reduced the likelihood of timely treatment for children under five, echoing findings from a study in Nigeria (16). caregivers’ fears of side effects from malaria drugs in children under five were linked to a decreased likelihood of seeking timely treatment for their under-fives, a result that mirrored the findings of another study conducted in Southwest Ethiopia (22).

The study revealed that caregivers who could recognize the early symptoms of malaria in their children were more likely to seek timely treatment, aligning with the expectation that well-informed caregivers may promptly identify signs and thus seek timely treatment. This finding corresponds with studies in Mali, Nigeria, and Madagascar, where caregivers’ ability to detect malaria symptoms in their children was linked to increased early treatment for under-five children

(18). The policymakers should implement health education programs for caregivers to improve their awareness and knowledge of recognizing early malaria symptoms in under-five children to improve timely malaria treatments.

In relation to their ability to pay, this study revealed that caregivers who possessed health insurance coverage were more likely to receive timely treatment for their under-five children with malaria. This finding aligns with expectations, as individuals with health insurance may be more health-conscious and informed, leading to a proactive approach in seeking prompt medical care for their children (35). This finding was consistent with a similar study conducted in mainland Equatorial Guinea, which also revealed that children whose caregivers lacked health insurance were significantly more prone to delayed malaria treatment (34). Healthcare authorities, community leaders, and health insurance providers should collaborate to create an affordable, context-specific insurance options to increase caregivers’ access to timely malaria treatment for under-five children.

This study also highlights that caregiver who sought help from herbalists or traditional healers before turning to formal healthcare facilities were less likely to get timely malaria treatment to their under-five children compared to those that did not get help from herbalist or traditional healers. This may suggest that these caregivers may initially opt for herbal or traditional remedies, possibly delaying healthcare facility visits until these alternative treatments are ineffective. The study reveals that around eighteen percent of caregivers resorted to herbal medicine before seeking treatment at healthcare facilities. This practice reflects a broader trend in Africa, where a significant proportion of the population, estimated at seventy to eighty percent, often turns to traditional herbalists or healers for treatment (36). This study’s findings are consistent with a study conducted in Nigeria(16). There is need for integration of traditional healers into the formal healthcare system, with training them to conduct Rapid Diagnostic Tests and administer antimalarials, to enhance timely malaria treatment for under-five children.

Finally, the concern over potential side effects of malaria drugs on under-five children was also identified as a significant factor affecting the timeliness of malaria treatment among under-fives, as indicated in this current study. Caregivers fearing malaria drug side effects were less likely to seek timely treatment for their children as compared to those that had no such fears. This may suggest that to they try alternative remedies before seeking formal healthcare, causing delays in treatment. This finding was similar to another study done in Southwest Ethiopia which also found that caregivers who had fear of the side effects of malaria drugs in their children were more likely to delay for malaria treatment for their under-five children (22). Malaria stakeholders should improve adherence to recommended treatments by dispelling misconceptions and addressing fears related to the side effects of malaria drugs.

Moreover, in relation to their ability to reach healthcare facilities, none of the factors were found to be significantly associated with timely malaria treatment of under five children in Kisumu East sub-county, Kenya. This discovery contradicted the results of a separate study conducted in in southern Malawi, which identified distance to health facilities as a significant determinant of timely malaria treatment (27). Additional another study in southeastern Nigeria supported the significant associations between physical factors and the prompt treatment of malaria (21). This finding may signify that in this context, geographical accessibility and transportation may not be the key determinants of timely malaria treatment as more caregivers reported short travel times and easy access to transportation, often choosing to walk to healthcare facilities. To validate this finding and gain a deeper understanding, further studies with larger sample sizes in similar contexts should be conducted.

This study’s strength lies in its use of the Conceptual Framework for Patient-Cantered Access to Health, offering a comprehensive view of healthcare access beyond facility entry. It provides valuable insights into determinants of timely malaria treatment in under-five children, aiding strategic planning for malaria control interventions at various levels. However, certain limitations must be considered. The hospital-based nature of the study could introduce selection bias as participating caregivers may not fully represent the general population. Additionally, the study’s findings rely on caregivers’ ability to accurately determine the onset of malaria symptoms (fever) in their under-five children. This subjective self-reporting may introduce recall bias or misreporting.

## Conclusion

Our study shows that ability to tell malaria symptoms, use of approved malaria drug, use of over-the-counter medication, having health insurance and seeking timely malaria care previously were associated with timely malaria treatment among under-five while herbalist visit, decision to seek care by the caregivers themselves, cost of malaria treatment and fear of malaria drug’s side effects were associated with delay of malaria treatment among under-five. Notably, caregivers who demonstrated the ability to identify symptoms of malaria, held belief of use of approved malaria drugs, turned to over-the-counter medication, possessed health insurance, and had prior experience of seeking timely malaria care exhibited a higher likelihood of timely malaria treatment for their sick under-five children while Caregivers who resorted to herbalist visits, made care decisions themselves, faced financial barriers, and harbored concerns about potential side effects of malaria drugs were more inclined to delayed malaria treatment for their under-five children.

## Data Availability

Data will be made available upon request

## DECLARATIONS

## Abbreviations

KIMS: Kenya Malaria Indicator Survey
KNBS: Kenya National Bureau of Statistics
RDT: Rapid Diagnostic Test
MOH: Ministry of Health Kenya
WHO: World Health Organisation
SDGs: Sustainable Development Goals
AOR: Adjusted Odds Ratio
PMI: President Malaria Initiative

## Acknowledgements

Geofrey Ochieng (GO) is a recipient of a TDR scholarship under the Postgraduate Training Scheme in Implementation Research at the University of Zambia. We are grateful to the financial support for the training scheme as provided by the UNICEF/UNDP/World Bank/WHO Special Program for Research and Training in Tropical Diseases (TDR). We are also grateful to the Ministry of Health in Kisumu County and the facility in-charges at the seven public health facilities where this research was conducted.

## Authors Contribution

GO and CJ was involved in conception and design of the study. Data analysis was done by GO. The first draft was done by GO. Review of the manuscript was done by CJ, JZ, AS, MS, PM and JK. All authors read, revised, and approved the final manuscript.

## Corresponding authors’ information

The main author, Geofrey Ochieng, can be contacted at the following email address. Geofrey Ochieng is a Masters student at the University of Zambia, in the School of Public Health within the Department of Epidemiology. His academic pursuits are focused on Epidemiology with a specialization in implementation science. If there are any inquiries or correspondence related to this research, please direct them to Geofrey Ochieng at the provided email address.

## Availability of data and materials

The dataset utilized and/or analyzed in this study are accessible and can be obtained from the corresponding author, Geofrey Ochieng, upon making a reasonable request.

## Consent for publication

Not applicable

## References

1. WHO. Global technical strategy for malaria 2016-2030, 2021 update [Internet]. World Health Organization. 2021. 1–40 p. Available from: https://apps.who.int/iris/rest/bitstreams/1357541/retrieve

2. WHO. World malaria report 2021 [Internet] [Internet]. World Health Organization. 2022. 2013–2015 p. Available from: https://www.who.int/teams/global-malaria-programme/reports/world-malaria-report-2021

3. WHO. Regional data and trends briefing kit World malaria. World Malar Rep 2022. 2022;(December):1–16.

4. KMIS. Kenya Malaria Indicator Survey 2020. 2021;158. Available from: www.nmcp.or.ke.

5. MoH. Kenya Malaria Programme Review 2018. 2019;

6. OAU. The Abuja Declaration on Roll Back Malaria in Africa. 2000;(April 2000):6–7.

7. USAID. PRESIDENT ’ S MALARIA INITIATIVE Kenya Malaria Operational Plan FY 2019. 2019.

8. MOH. Towards a Malaria-Free Kenya: Kenya Malaria Strategy 2019. 2019;1–88.

9. Marsh VM, Mutemi WM, Willetts A, Bayah K, Were S, Ross A, et al. Improving malaria home treatment by training drug retailers in rural Kenya. Vol. 9, Tropical Medicine and International Health. 2004. p. 451–60.

10. Chuma J, Musimbi J, Okungu V, Goodman C, Molyneux C. Reducing user fees for primary health care in Kenya: Policy on paper or policy in practice? Int J Equity Health. 2009;8:1–10.

11. Mwenesi HA. Social science research in malaria prevention, management and control in the last two decades: An overview. Acta Trop. 2005;95(3):292–7.

12. Wiseman V, Scott A, Conteh L, McElroy B, Stevens W. Determinants of provider choice for malaria treatment: Experiences from The Gambia. Soc Sci Med. 2008;67(4):487–96.

13. Mburu CM, Bukachi SA, Shilabukha K, Tokpa KH, Ezekiel M, Fokou G. Determinants of treatment-seeking behavior during self-reported febrile illness episodes using the socio-ecological model in Kilombero District , Tanzania. BMC Public Health. 2021;1–11.

14. Lu G, Cao Y, Chai L, Li S, Heuschen AK, Chen Q, et al. Barriers to seeking health care among returning travellers with malaria_ A systematic review. Trop Med Int Heal. 2021;

15. Shiferaw BD, Geremew M, Kumalo A. Determinants of Delay in Malaria Treatment Seeking for Under-Five Children with Malaria Attending Health Centers of Bench-Maji Zone , South-Western Ethiopia Case-Control Study DeterminantsofDelayinMalariaTreatmentSeekingforUnderFiveChildrenwithMalariaAt. 2018;(January).

16. Eseigbe EE, Anyiam JO, Ogunrinde GO, Wammanda RD, Zoaka HA. Health Care Seeking Behavior among Caregivers of Sick Children Who Had Cerebral Malaria in Northwestern Nigeria. Malar J. 2012;2012 (December 2009).

17. Xu JW, Xu QZ, Liu H, Zeng YR. Malaria treatment-seeking behaviour and related factors of Wa ethnic minority in Myanmar: A cross-sectional study. Malar J. 2012;

18. Do M, Babalola S, Awantang G, Toso M, Lewicky N, Pompsett A. Associations between malaria-related ideational factors and care-seeking behavior for fever among children under five in Mali, Nigeria, and Madagascar _ Enhanced Reader.pdf. PLoS One. 2018;

19. McCombie SC. TREATMENT SEEKING FOR MALARIA : A REVIEW OF RECENT RESEARCH *. 1996;43(6):933–45.

20. Tesfahunegn A, Zenebe D, Addisu A. Determinants of malaria treatment delay in northwestern zone of Tigray region, Northern Ethiopia, 2018. Malar J [Internet]. 2019;18(1):358. Available from: 10.1186/s12936-019-2992-7

21. M. Chukwuocha U, Iwuoha GN, Nwakwuo GC, Egbe K, Ezeihekaibe CD, Ekiyor CP, et al. Malaria care-seeking behaviour among HIV-infected patients receiving antiretroviral treatment in South-Eastern Nigeria : A cross-sectional study. PLoS One. 2019;1–16.

22. Shumerga AT, Hebo HJ, Gebrehiwot TT, Gebre MN. Determinants of Delay in Seeking Malaria Treatment for Under-Five Children at Gambella Town , Southwest Ethiopia : A Case-Control Study. 2020;2020.

23. Mitiku I, Assefa A. Caregivers’ perception of malaria and treatment-seeking behaviour for under five children in Mandura District, West Ethiopia: A cross-sectional study. Malar J. 2017;16(1):1–10.

24. Getahun A, Deribe K, Deribew A. Determinants of delay in malaria treatment-seeking behaviour for under-five children in south-west Ethiopia : a case control study. 2010;1–6.

25. Shah JA, Emina JBO, Eckert E, Ye Y. Prompt access to effective malaria treatment among children under five in sub-Saharan Africa: A multi-country analysis of national household survey data. Malar J. 2015;14(1):1–14.

26. Moise J, Kaboré T, Siribié M, Hien D, Soulama I, Barry N, et al. Attitudes , practices , and determinants of community care - seeking behaviours for fever / malaria episodes in the context of the implementation of multiple first - line therapies for uncomplicated malaria in the health district of Kaya , Burkina Faso. Malar J [Internet]. 2022;1–14. Available from: 10.1186/s12936-022-04180-z

27. Masangwi SJ, Ferguson NS, Grimason AM, Morse TD, Kazembe LN, Masangwi SJ, et al. Care-seeking behaviour and implications for malaria control in southern Malawi Care-seeking behaviour and implications for malaria control in southern Malawi. South African J Epidemiol Infect. 2017;8782(December).

28. Chuma J, Okungu V, Molyneux C. Barriers to prompt and effective malaria treatment among the poorest population in Kenya. 2010;1–14.

29. Levesque JF, Harris MF, Russell G. Patient-centred access to health care: conceptualising access at the interface of health systems and populations. Int J Public Euiquity Heal. 2013;28(3):3262–5.

30. KNBS. 2019 Kenya Population and Housing Census [Internet]. Vol. II, 2019 Kenya Population and Housing Census. 2019. 251 p. Available from: http://www.knbs.or.ke

31. Workineh B, Mekonnen FA. Early treatment - seeking behaviour for malaria in febrile patients in northwest. Malar J [Internet]. 2018;1–8. Available from: 10.1186/s12936-018-2556-2

32. Tiruneh M, Gebregergs GB, Birhanu D. Determinants of delay in seeking treatment among malaria patients in Dera district , NorthWest Ethiopia : a case control study. 2018;18(3):552–9.

33. Jayatillake R V, Sooriyarachchi MR, Senarathna DLP. Adjusting for a cluster effect in the logistic regression model : an illustration of theory and its application. 2011;39(3):211–8.

34. Romay-Barja M, Cano J, Ncogo P, Nseng G, Santana-Morales MA, Valladares B, et al. Determinants of delay in malaria care-seeking behaviour for children 15 years and under in Bata district, Equatorial Guinea. Malar J. 2016;15(1):1–8.

35. Al-Hanawi MK, Mwale ML, Kamninga TM. The effects of health insurance on health-seeking behaviour: Evidence from the Kingdom of Saudi Arabia. Risk Manag Healthc Policy. 2020;13:595–607.

36. Coker HAB, Adesegun S. The malaria scourge: The place of complementary traditional medicine. Niger Med Pract. 2006;5(2):126–32.

